# Temporary Immunity and Multiple Waves of COVID-19

**DOI:** 10.1101/2020.07.01.20144394

**Authors:** B Shayak, Mohit M Sharma, Anoop Misra

## Abstract

In this work we use mathematical modeling to describe the potential phenomena which may occur if immunity to COVID-19 lasts for a finite time instead of being permanent, i.e. if a recovered COVID-19 patient may again become susceptible to the virus after a given time interval following his/her recovery. Whether this really happens or not is unknown at the current time. If it does happen, then we find that for certain combinations of parameter values (social mobility, contact tracing, immunity threshold duration etc), the disease can keep recurring in wave after wave of outbreaks, with a periodicity approximately equal to twice the immunity threshold. Such cyclical attacks can be prevented trivially if public health interventions are strong enough to contain the disease outright. Of greater interest is the finding that should such effective interventions not prove possible, then also the second and subsequent waves can be forestalled by a consciously relaxed intervention level which finishes off the first wave before the immunity threshold is breached. Such an approach leads to higher case counts in the immediate term but significantly lower counts in the long term as well as a drastically shortened overall course of the epidemic.

As we write this, there are more than 1,00,00,000 cases (at least, detected cases) and more than 5,00,000 deaths due to COVID-19 all over the globe. The unknowns surrounding this disease outnumber the knowns by orders of magnitude. One of these unknowns is how long does immunity last i.e., once a person recovers from COVID-19 infection, how long does s/he remain insusceptible to a fresh infection. Most modeling studies assume lifetime immunity, or at least sufficiently prolonged immunity as to last until the outbreak is completely over. Among the exceptions are Giordano et. al. [1] and Bjornstad et. al. [2] who account for the possibility of re-infection – while the former find no special behaviour on account of this, the latter find an oscillatory approach towards the eventual equilibrium. In an article which appeared today, Kosinski [3] has found multiple waves of COVID-19 if the immunity threshold is finite. The question of whether COVID-19 re-infection can occur is completely open as of now. A study [4] has found that for benign coronaviruses (NOT the COVID-19 pathogen!), antibodies become significantly weaker six months after the original infection, and re-infection is common from one year onwards. Although it is currently unknown whether COVID-19 re-infections can occur, the mere possibility is sufficiently frightening as to warrant a discussion of what might happen if it is true. In this Article, we use mathematical modeling to present such a discussion. Before starting off, **let us declare in the clearest possible terms that this entire Article is a what-if analysis, predicated on an assumption whose veracity is not known at the current time**. The contents of this Article are therefore hypothetical – as of now they are neither factual nor counter-factual.

## §1 EQUATION

We start from the retarded logistic equation which we have derived in a recent work [5] as a universal model for the spread of COVID-19. If *y* (*t*) is the cumulative number of corona cases in a region and *N* the region’s total susceptible population, then this equation is

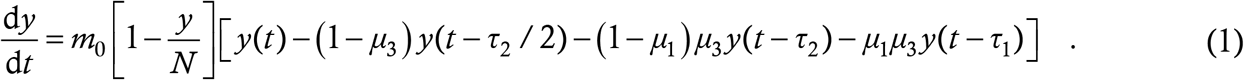

The parameters here are

- *m*_0_: the rate (people/day) at which a corona case at large in society spreads the disease to healthy people at the start of the epidemic
- *τ*_1_: the asymptomatic infection period which we take to be 7 days throughout
- *τ*_2_: the latency period during which a symptomatic patient can transmit the disease prior to manifesting symptoms. We take it to be 3 days throughout
- *μ*_1_: a number between 0 and 1, it denotes the fraction of patients who are asymptomatic
- *μ*_3_: a number between 0 and 1, it denotes the fraction of patients (both symptomatic and asymptomatic) who are NOT detected in contact tracing drives and quarantined

We shall not go into the details of the derivation but refer to Ref. [5] for that. Very quickly, the right hand side (RHS) of (1) has three terms whose significance is as follows. The first term is *m*_0_. It is the product of two things – the rate at which the case at large interacts with other people, and the probability that every interaction results in a transmission. *m*_0_ is defined at the start of the outbreak when everyone is susceptible – intrinsic insusceptibles are excluded at the outset. The net value of *m*_0_ is found by averaging over all at large cases. The second term on the RHS is the logistic term 1 − *y*/*N*. This represents the probability that the person whom the at large case is interacting with really is susceptible. In Ref. [5] we had assumed everlasting immunity, so the probability of a random person’s being a recovered case (and hence insusceptible) was *y*/*N* and the probability of his/her not being a case (and hence susceptible) was 1 − *y*/*N*. (There are three reasons why we used *y* in the numerator rather than a delayed *y* which accounts for the finite recovery period: (*a*) we assumed that at any instant the difference between the total and recovered case counts is small, (*b*) we wished to retain mathematical simplicity of the model, and (*c*) while retaining simplicity, we also wished to explicitly build in the very important feature that 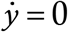 if *y* = *N*.) This logistic term is the thing which will have to be modified here. Before that, the third term on the RHS of (1) is the big box bracket which accounts for undetected asymptomatics remaining at large for time *τ*_1_, undetected latent patients remaining free for time *τ*_2_ and the rest. This term remains unchanged in this Article.

The modification to the logistic term is not difficult to implement since the philosophy remains unchanged; the difference lies only in the details (just to be clear, the solutions we’ll get here will NOT differ from Ref. [5] only in details). The term must still denote the probability that a random person is susceptible. Now, we assume that each recovered case remains immune or insusceptible for a duration *τ*_0_, the immunity threshold, typically of the order of weeks or months. So, if a person once contracts and recovers from the infection at time *t*, then s/he remains immune upto time *t* + *τ*_0_ and then becomes susceptible again. At any given instant, the people who are immune are all those who have contracted the infection during the last *τ*_0_ days, and no one else. Hence, the number of insusceptibles at time *t* is exactly the number of new infections which have occurred between time *t* − *τ*_0_ and *t*, which is *y* (*t*) − *y* (*t* − *τ*_0_). The structure of this term and the underlying logic are the same as those motivating the other delay terms in (1); please consult Refs. [5,6] for more details of the derivation. In proposing this structure, we have again assumed that recoveries are instantaneous (see the previous paragraph) and also have ignored deaths. Since the mortality rate of COVID-19 is fortunately quite low, this second assumption is reasonable as well. Then, the probability that a random person at time *t* is susceptible is

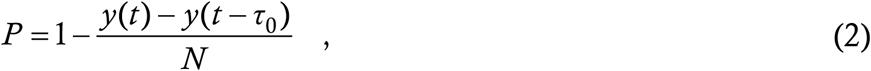

and working this into (1) yields

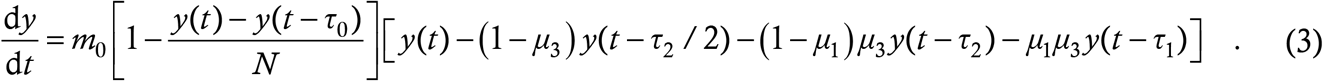

This is the model equation which we shall use to describe the spread of Coronavirus with immunity threshold *τ*_0_. The philosophy of the model is similar to the SEIS or SEIRS models [2,3,7,8,9]; the structure however is novel.

Just as in Ref. [5], *y* = const. is a solution of (3) for all constants, since it makes the big long box bracket vanish. A second condition under which 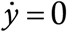 is if *y* (*t*) − *y* (*t* − *τ*_0_) = *N*. If *t* = 0 denotes the start of the epidemic, then *y* is automatically zero for all *t* < 0; the above condition then holds if *y* (*t*) becomes equal to *N* at any time before and upto *t* = *τ*_0_. This is a physically transparent condition – it means that if everyone gets infected before the first patient loses immunity, then the epidemic automatically ceases. These are plausibility checks of our model (3) – we now go on to the solution classes.

## §2 SOLUTIONS

**Disclaimer: All results in this Section are based on an arbitrarily assumed immunity threshold of 200 days. There is currently no evidence either to indicate that such a threshold exists for the COVID-19 virus or to estimate its value**.

In Ref. [5] we have calculated the reproduction number *R* of (1) by assuming *y* to be a constant. To calculate *R* from (3), we must assume that not only *y* but also *y* (*t* − *τ*_0_) is constant. If we make this assumption, then we get:

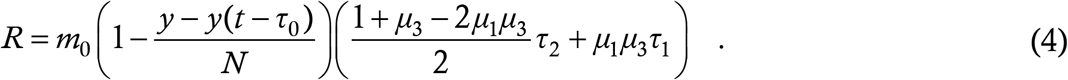

The extent to which this expression is meaningful in the present situation remains to be analysed. What is true though is that, given all the parameter values, *R* has a maximum value when *y* − *y* (*t* − *τ*_0_) = 0, which holds for example at the start of the outbreak; this maximum is *R*_0_. It is still valid in the present situation that if *R*_0_ < 1 then there is no spread of the disease in any case.

Before presenting the solutions of (3), we first gain intuition through some fictitious pedagogical examples. We had introduced one such example in Ref. [6], which we recapitulate briefly. This example was the fantasy kingdom of Coronaland where the king declares a 28-day “100 percent lockdown” – literally nobody is allowed to step out of his/her house. Because Coronaland is fictitious, this doesn’t pose a logistical problem of survival. At the end of the lockdown, every case has either recovered or died at home, and the virus doesn’t exist any longer when normal life is restarted. This example remains valid for our present purposes.

Our next example is the city of Coronapur. This has an initial population of 500 susceptible people, and has no cases to start with. The immunity threshold is 200 days and every case recovers within a day (consistent with the modeling assumptions above). The spread of the virus here is governed by the following rule:

- On any day if there are 5 or more than 5 susceptible individuals present, then exactly 5 of them (randomly selected where necessary) contract the virus
- On any day if there are less than 5 susceptible individuals present then all of them contract the virus

The case trajectory of Coronapur is simple enough to predict. There are total 5 cases on day 1, 10 on day 2, 15 on day 3 and so on, all the way upto 500 on day 100. Then, since the first batch of cases is still well within its immunity period, there are no more susceptible people left and the propagation stops. On day 100, the last batch of cases also clears the virus which then doesn’t exist in Coronapur any longer. Everyone has been infected but the virus is dead and the epidemic over.

The city of Coronabad has everything same as Coronapur except that the number 5 is replaced by 2. The case trajectory starts off like a 2 times-table – 2 cases on the first day, 4 on the second, 6 on the third, 200 on the 100^th^ (unlike the 500 of Coronapur) and so on upto 400 cases on the 200^th^ day. Thus, on day 201, there are 100 people who are yet to contract the infection. But, on this day, the cases of day 1 lose their immunity and also get added to the susceptible pool. So the two new infections of day 201 can be anyone from this pool of 102. Similarly on day 202, the two cases of day 201 get removed from the susceptible pool but the two recoveries from day 2 get added and the pool size doesn’t change. This scenario remains invariant for all time – every day there is a susceptible pool of 102 and every day there are 2 new infections from amongst this pool. Even though the initial growth rate is much slower than in Coronapur, the city of Coronabad becomes a land of immortal virus.

Now we present the solutions of (3). While perturbation theoretic inroads into this nonlinear delay differential equation (DDE) will be a fascinating exercise, they will not be of too much utility in actually forecasting the spread of corona. Hence we leave this study until later, and solve the equations numerically using the routine of Refs. [5,6] – 2^nd^ order Runge Kutta with step size 0.001 day. A DDE needs to be seeded with an initial function of duration equal to the maximum delay in the problem. This is the delay *τ*_0_, which is 200 days. The seeding function we take is zero cases for the first 193 days and then linear growth of cases at 100 cases/day for the next seven days. We take *t* = 0 to be the time when the case count first departs from zero i.e. the 194^th^ day of the seeding period.

There is only one issue which needs special mention. This is that, in a numerical simulation, the case rate will never be identically zero but will be something like 0.001 cases/day (or machine epsilon cases/day). This can pose a serious problem in a situation where there are potential second waves of epidemic. To circumvent this issue, we have arranged for manual termination of the run if the case rate becomes low enough. To make the termination condition more precise, we define the number of active cases at time *t* to be *y* (*t*) − *y* (*t* − 14) and stop the run if there is less than one active case for 14 consecutive days. While the number 14 (twice) is somewhat arbitrary, the criterion of a low enough active case count for a long enough period is a very reasonable indicator of the true end of the outbreak. We run all simulations either upto *t* = 1400 days or until they terminate, whichever is earlier.

To present the solution classes of (3), we use the same Notional Cities as we did in Ref. [5]. Each city has an initial susceptible population of *N* = 300000 (3 lakhs) and has 80 percent asymptomatic carriers i.e. *μ*_1_ = 0.8. City A gets an A for public health – it has *m*_0_ = 0.23 and *μ*_3_ = 1/2, corresponding to good contact tracing in a hard lockdown. With *R*_0_ < 1, in Ref. [5] it burnt out the epidemic in about 120 days – let’s see what it does here. Just as in there, we show three things in the same plot – the function *y* (*t*) as a blue line, its derivative 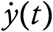 as a green line and the “epi-curve” or weekly increments in cases, scaled down by a factor of 7, in grey bars.

This was only to be expected – City A is the realistic version of Coronaland, like New Zealand. It will make no difference for New Zealanders whether immunity to COVID-19 is permanent or temporary.

Next, we break alphabetical order and present City C. This is the city which has less effective contact tracing and keeps itself open throughout the epidemic – *μ*_3_ = 3/4 and *m*_0_ = 0.5 all the time. (We use 0.23 for a “low” value of *m*_0_ since that number came out of data fits in Ref. [5]; the “high” value of 0.5 is just designed to generate 90 percent infection level at the end of the outbreak.) In Ref. [5], City C exploded with more than 2.5 lakh infections over just 50 days.

Here, it does the same. This city is the equivalent of conceptual Coronapur – everyone gets infected quickly and after that the virus is dead.

Now we come to City B. This implements a lockdown but has less effective contact tracing – *m*_0_ = 0.23 like City A but *μ*_3_ = 3/4 like City C. In Ref. [5], City B was the next best to A, crawling up to 80,000 infections over a period of 220 days and judiciously avoiding overstressing of the healthcare facilities. Here however, that 220-day run becomes longer than the immunity threshold, so we might be in for some fun.

This is indeed the real-world version of Coronabad – the growth rate at any instant is low but the flipside is wave after wave of outbreaks. Now, we are not fool enough to believe that corona can really go on for four years like we have shown – by that time there will surely be a cure, a vaccine or (the likeliest possibility?) a harmless mutation which will make the disease go away. The very long simulation runtime is just to show the periodicity of the case trajectories – it seems that the period is very nearly equal to 400 days or 2*τ*_0_. Why this is so is a question we leave for later. A second wave coming about a year after the first is however a realistic possibility; it is very much in the public mind and is something for which we should be prepared. The second and subsequent waves which we have obtained here are “natural” waves in the sense that they are brought about by intrinsic biological properties of the virus and the human immune system – they are not “artificial” waves precipitated by periodic variation of the strength of intervention measures. Another thing to note is that the case count at the end of the run is greater than the city’s initial susceptible population, so at least some people have been infected at least twice.

City D of Ref. [5] was a late lockdowner – with *μ*_3_ = 3/4 throughout, it kept *m*_0_ = 0.5 until 40,000 cases and then cut down to *m*_0_ = 0.23 for the rest of the outbreak. The ultimate result was termination of the epidemic at 70 days and 1,38,000 cases. This duration is short enough for immunity breach not to occur, so we expect the plot to remain unchanged here. Indeed it does and we dispense with the repetition. Cities E and F of Ref. [5] were reopeners. Like City A, City E ensured *R* < 1 throughout via skilful public health measures and extinguished the outbreak in time. Once again, it remains the same here and we don’t show it.

City F is more interesting. In Ref. [5] it started under conditions of burnout but increased mobility abruptly at 80 days, resulting in a (now, artificial) second wave of infections. Here, we keep the philosophy the same but change the parameters a bit. This time, we start off F with the parameter values of B, on a path to viral immortality. 150 days into the outbreak however, it gets wind of the finite immunity threshold (probably from another city which has suffered the outbreak earlier in time). Since City F lacks the wherewithal to drive the epidemic to self-burnout, it adopts a different strategy. It reopens completely, raising *m*_0_ to 0.5, aiming to throw its entire population to the virus before immunity runs out. Here is the plot.

Howsoever controversial this strategy might appear, it works. The reopening generates an artificial second wave and very likely overstresses healthcare systems after the 150^th^ day, but it does succeed in making the epidemic vanish completely at 210 days and 2,46,000 infections. Not only is the case count lower than in City B but the economic and psychological gains relative to that city are immeasurable.

City G is a variant of F. Like F, it gets wind of the immunity threshold and reopens on the 150^th^ day, trying to beat the second wave. Unlike F however, it fears for its healthcare system and opts for some restraint during the reopening – it goes for *m*_0_ = 0.4 instead of 0.5.

While the rate at the height of the artificial second wave is about half of F’s, that reduction comes at the expense of a more spread out peak. This ensures that the epidemic isn’t dead but merely smouldering by the time the initial recoveries start losing immunity. When the susceptible pool becomes large enough, the glowing embers start a conflagration – a natural second wave of gigantic proportions, as it turns out. Only when this wave has torn through the entire population does the epidemic become history. This catastrophe can be averted however if the higher mobility of 0.4 is initiated earlier in the first wave, even by a mere 10 days it turns out (we don’t show the plot again).

Now we present a couple of plots of a more mathematical character. We consider City B, which displayed the wave solution. For *μ*_3_ = 3/4, the critical value of *m*_0_ which results in *R*_0_ just crossing unity happens to be 0.199. If we start things off with *m*_0_ = 0.19, the solution burns out in time like City A, as expected. For *m*_0_ = 0.20, here’s what we have (now onwards, we are letting go of the epi-curve).

This value of *m*_0_ is a hairline above the stability threshold, but in the infinite-immunity case [5], *R* dips below 1 at only a few hundred infections, from which point onwards the epidemic burns itself out. This initial behaviour – a back-bending curve – is seen here also. Then however, after crossing the immunity threshold, the solution becomes linear in time and this linear solution is stable. When we increase *m*_0_ to 0.21, the following happens.

This time, there is a decaying oscillation superposed on a linearly growing solution – the growth rate is faster than in the previous case. Raising *m*_0_ to 0.22 produces a steady, large-amplitude oscillation superposed on linear growth, just as for City B in Fig. 3. This class of solution continues upto *m*_0_ = 0.25 when it gets replaced by a one-shot “logistic-like” solution of City C, Fig. 2. Perhaps the stable bounded solution for *R*_0_ < 1 first loses stability to a linear solution through a saddle-node bifurcation, which then undergoes a Hopf bifuraction to generate a periodic solution. This is only a conjecture. We suspect that we are finding the wave solutions in a narrow region of *m*_0_-space since only these few values of *m*_0_ in the logistic DDE (1) produce solutions which last as long as the immunity threshold. Reducing the threshold should increase the range of *m*_0_ values which produce the wave solution. The details of the bifuraction structure of (3) are not really relevant for predicting and dealing with corona – what they show us is that the mathematical analysis of disease DDEs like (1) and (3) can be extremely rewarding on its own.

**Figure 1:**
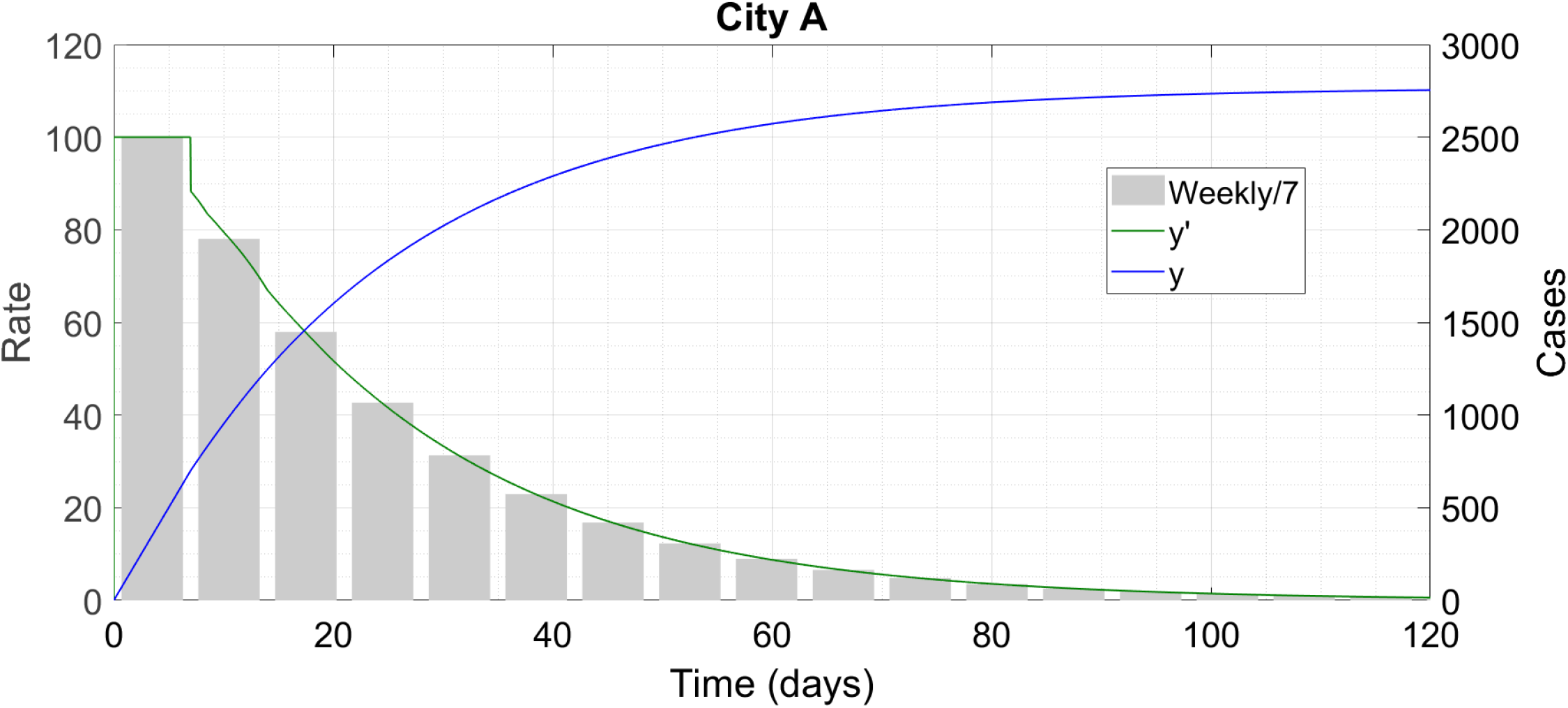
City A proceeds to self-burnout.

**Figure 2:**
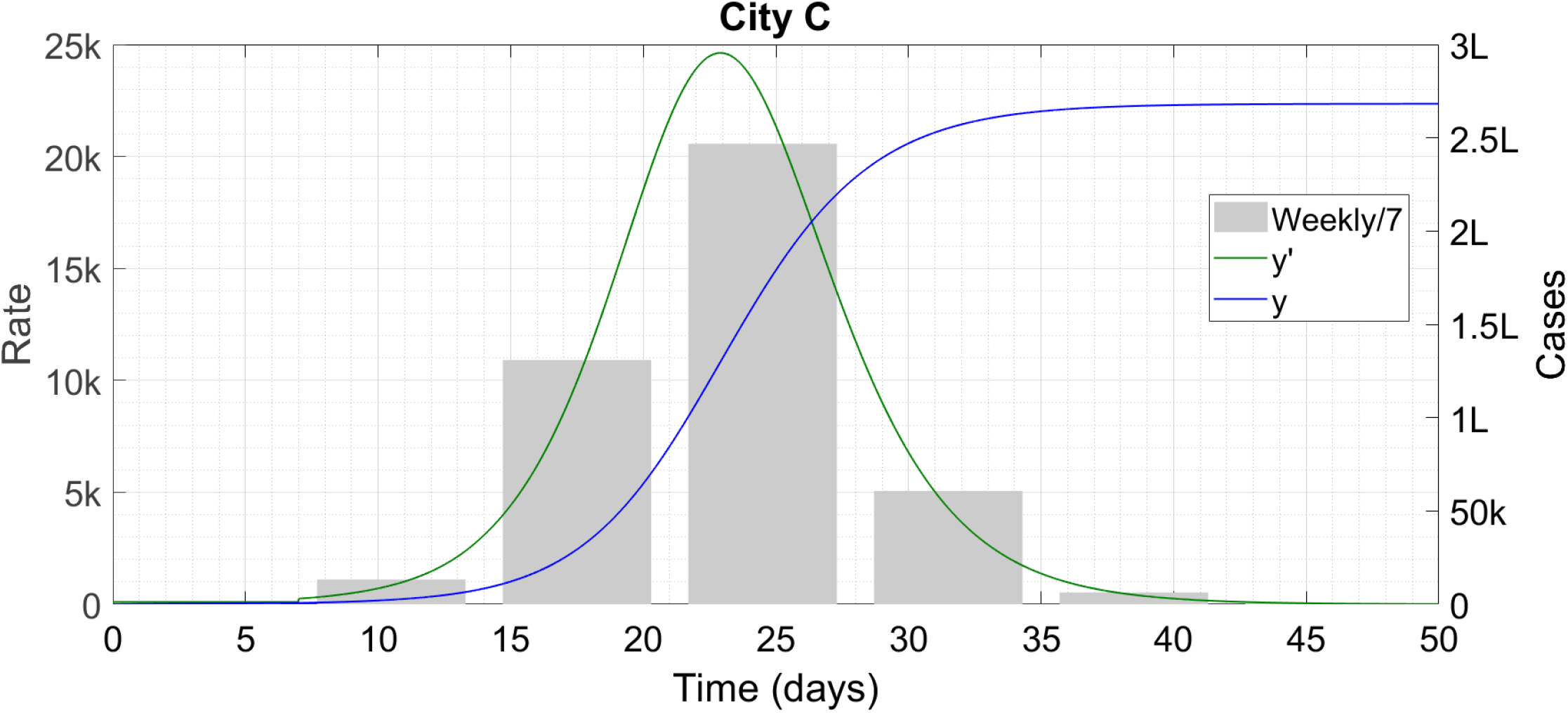
City C proceeds to herd immunity well within the 200-day period. ‘k’ denotes thousand and ‘L’ lakh.

**Figure 3:**
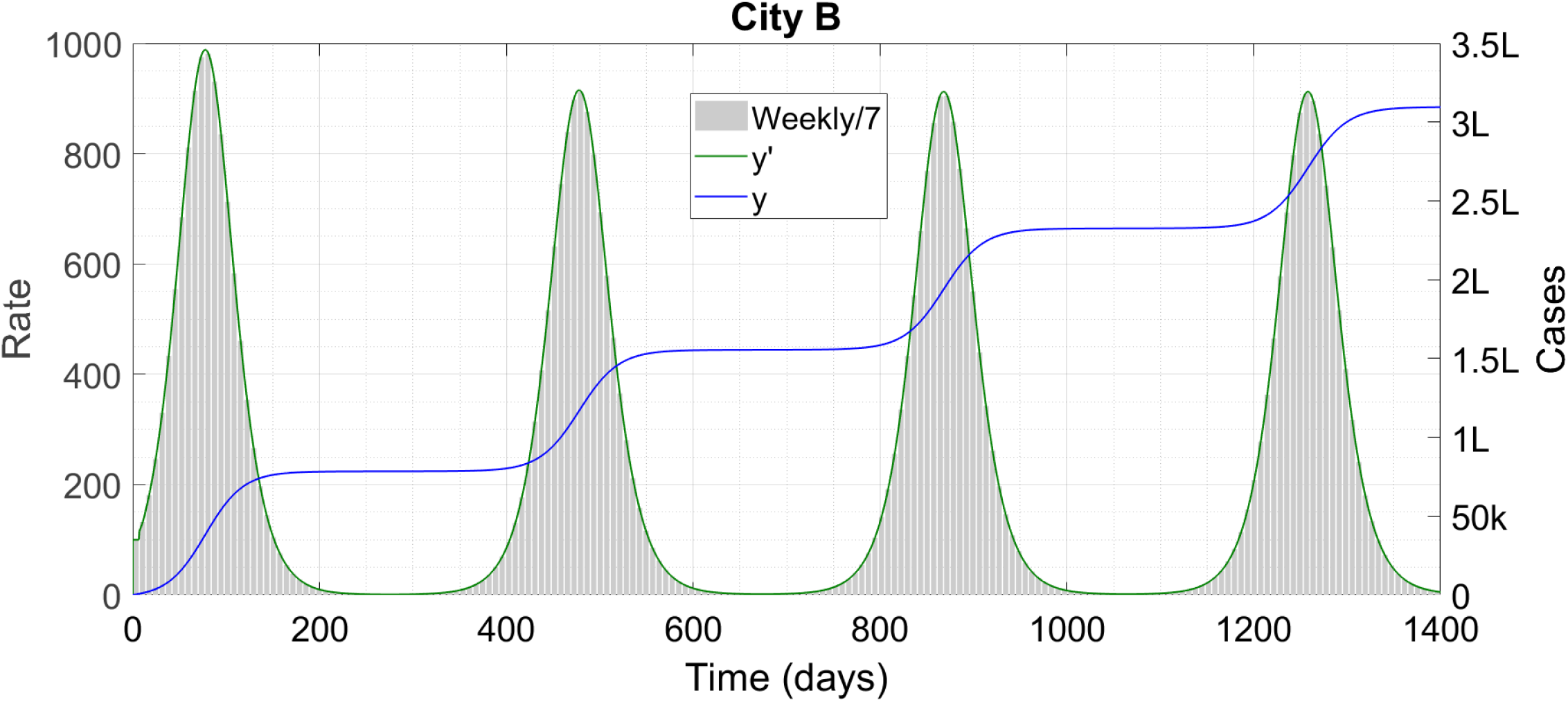
City B has multiple waves of COVID-19. ‘k’ denotes thousand and ‘L’ lakh. We have stopped the simulation at 1400 days – the waves go on after that time as well.

**Figure 4:**
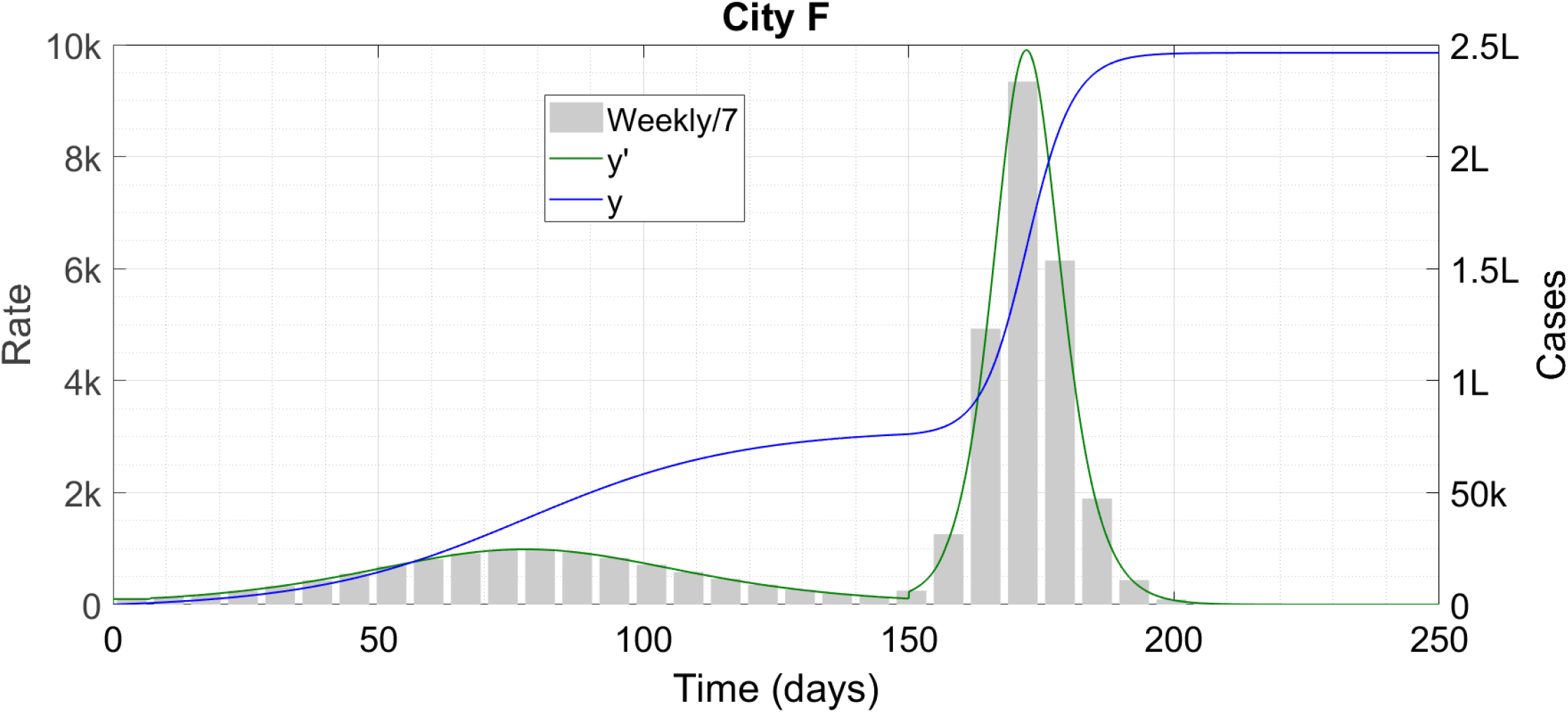
City F successfully pulls off a counter-intuitive epidemic management strategy. ‘k’ denotes thousand and ‘L’ lakh.

**Figure 5:**
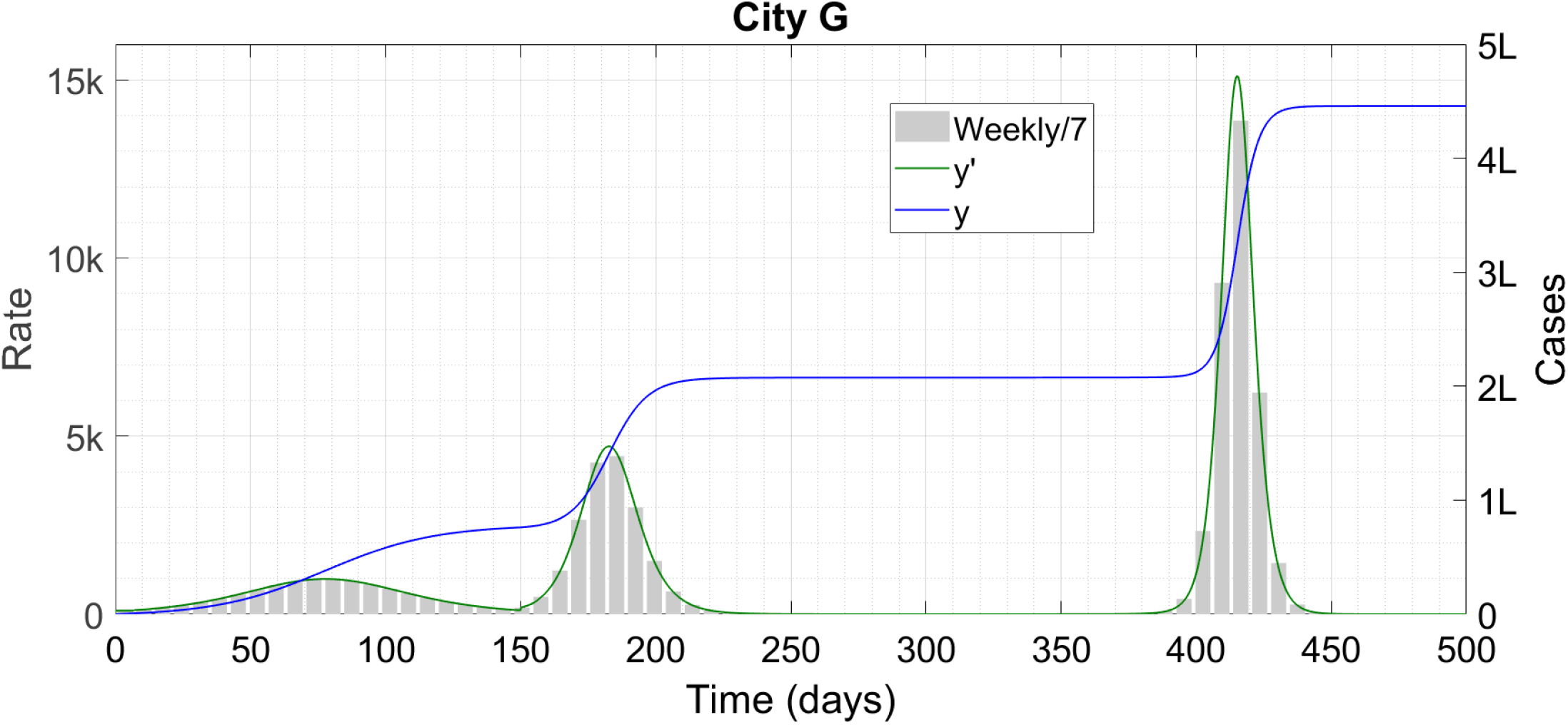
City G attempts the same strategy as F but it backfires. ‘k’ denotes thousand and ‘L’ lakh.

**Figure 6:**
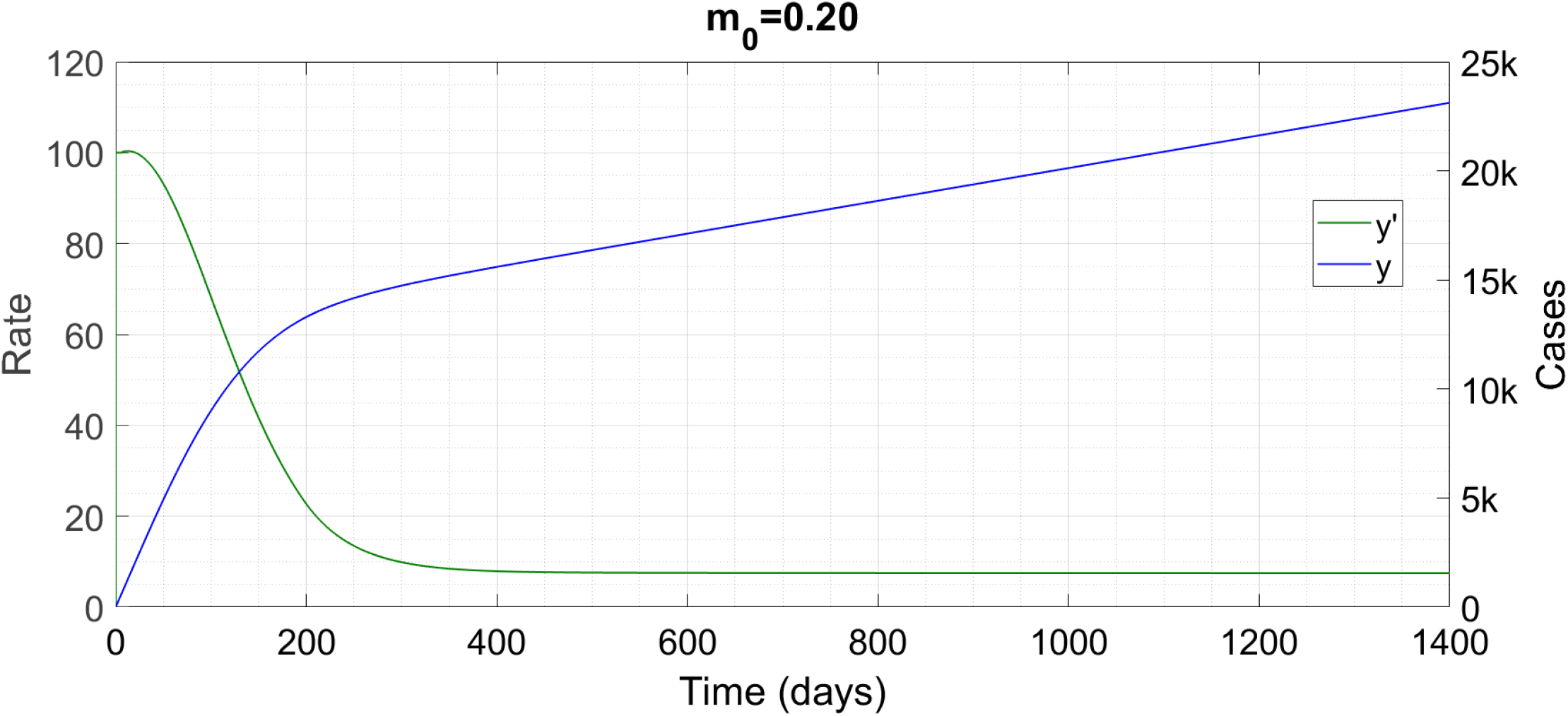
An R_0_ just above the critical leads to a linearly growing solution. ‘k’ denotes thousand.

**Figure 7:**
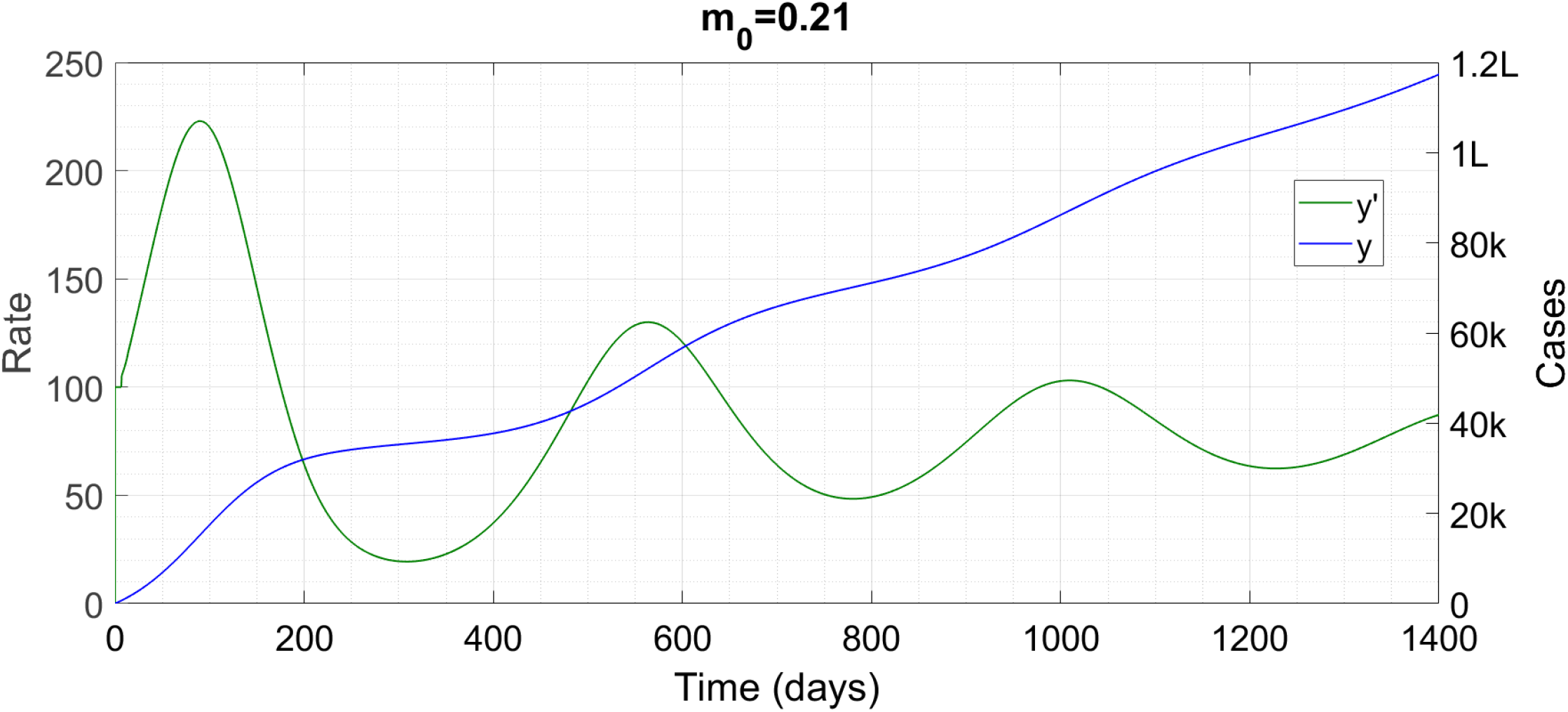
A slight increase in m_0_ adds a decaying oscillation to the linear solution. ‘k’ denotes thousand and ‘L’ lakh.

As an aside we note that much of the Literature [1,2,7,8,9] appears to have missed the multiple wave solutions possible with temporary immunity. References [1,8,9] find no special feature whatsoever with respect to the conventional SEIR models (infinite immunity). Reference [2] finds an oscillatory approach to an endemic equilibrium, while Ref. [6] finds steady oscillations about such an equilibrium – both features being absent in conventional models. The latter [6] is closer to our solution since the epidemic there also doesn’t die out in time. The Reference whose findings are most similar to ours is Ref. [3], since that work also gets the multiple waves. However, Ref. [3] has these waves whatever the value of *R*_0_, while we find them only for a range of *R*_0_, which is possibly even more realistic. We leave a more exhaustive Literature survey and comparison for the future.

## §3 IMPLICATIONS AND CONCLUSION

Before presenting our interpretation of the solutions, we again repeat the disclaimer that **the entire modeling study, and any conclusion drawn therefrom, is hypothetical and is predicated on a statement of currently unknown factual validity**. In practice, if an immunity threshold for COVID-19 does exist, we’ll first get it to know of it from those countries which have suffered their initial outbreak at an early stage and can be relied on to tell the truth. Then, non-burnout regions which have started off later will be in positions similar to the cities B to G. Those which have implemented delayed lockdowns, like New York City, USA, will be at an intrinsic advantage like City D. Those which have locked down early but reopened due to economic considerations, like many cities in India and USA, will be in positions similar to F and G. Using modeling, they might be able to predict their standing by the time immunity starts running out, and accordingly reopen further if need be. We also note that the disaster of City G is by and large an avoidable one since G does have a protracted period of extreme quiescence before the second wave begins. During this period, it should be possible to reimpose lockdown if necessary and perform the contact tracing and isolation required to stamp the epidemic out before the natural second wave begins.

In conclusion, what our Article does more than anything else is to add to the body of unknowns about this new and dangerous virus. Thanks to the extreme parameter sensitivity in the retarded logistic equation (1), the foundations of any public health measure are insecure to begin with (see §13 of Ref. [5]). This new possibility, and the extreme subtlety and variability of the resulting behaviours, makes things even more of a game of chance. Ultimately, the management of COVID-19, at least in most of the world, seems to be a problem too complex for us humans to handle, and we turn it over to the Ultimate Problem Creator-cum-Solver in the humble hope for an expedite solution.

## Data Availability

The manuscript uses NO data.

## Notes

### Competing Interest Statement

The authors have declared no competing interest.

### Funding Statement

We have NOT received any funding for this study from any agency.

### Author Declarations

No ethical guidelines were necessary due to purely mathematical modeling study. No IRB approval sought.

